# GWAS of cataract in Puerto Ricans identifies a novel large-effect variant in ITGA6

**DOI:** 10.1101/2023.07.25.23293173

**Authors:** Jingchunzi Shi, Jared O’Connell, Barry Hicks, Wei Wang, Katarzyna Bryc, Jennifer J. Brady, Vladimir Vacic, Will Freyman, Noura S. Abul-Husn, Adam Auton, 23andMe Research Team, Suyash Shringarpure

## Abstract

Cataract is a common cause of vision loss and affects millions of people worldwide. Genome-wide association studies (GWAS) and family studies of cataract have demonstrated a role for genetics in cataract susceptibility. However, most of these studies have been conducted in populations of European or Asian descent, leaving the genetic etiology of cataract among Hispanic/Latino (HL) populations unclear. Here we perform the first GWAS of cataract in a Puerto Rican population of research participants derived from the customer base of 23andMe, Inc. In our analysis with 3,060 self-reported cases and 41,890 controls, we found a novel association of large effect size with a rare coding variant in the ITGA6 gene (rs200560853, p-value=2.9×10^−12^, OR=12.7, 95% CI=[6.5, 24.7]). ITGA6 is part of the integrin alpha chain in the laminin receptor subfamily, and likely contributes to eye lens homeostasis, transparency, and cell survival. We found that this coding variant is associated with a 13.7 year earlier disease onset on average, as well as a 4.3-fold higher rate of cataract events in the Puerto Rican population. The variant has a minor allele frequency (MAF) of 0.089% in Puerto Rico and is extremely rare elsewhere in the world. Population genetic analyses showed that the variant is only found in individuals with ancestry from the Americas and countries bordering the Mediterranean Sea, suggesting a North African origin. Our discovery identifies a novel genetic risk factor for cataract in Puerto Ricans and highlights the importance of including underrepresented populations in genomics research to improve our understanding of disease in all populations.

## Introduction

Cataract is one of the leading causes of visual impairment, affecting 94 million people worldwide (World Health Organization: Global data on visual impairment). It manifests as a loss of transparency or clouding in the eye lens due to tissue degradation and protein clumping, resulting in progressive vision loss. The symptoms of cataract include blurred or hazy vision, reduced color intensity, increased sensitivity to glare, difficulty seeing at night, and changes in refractive error (National Eye Institute: Cataracts). Cataracts typically occur as an age-related disease, but may also be inherited with an early onset in association with DNA or protein damage which affects lens proteins and aquaporins (Wasnik, H. *et al*., 2021). Other risk factors include diabetes, certain medications, smoking, alcohol consumption, exposure to ultraviolet radiation, and nutritional deficiencies (National Eye Institute: Cataracts). The heritability of cataract is estimated to be between 35-58% based on twin and family studies (Hammond, C. J. *et al*., 2000; Hammond, C. J. *et al*., 2001; Congdon, N. *et al*., 2004; Sanfilippo, P. G. *et al*., 2010). A recent multi-ethnic genome-wide association study (GWAS) for cataract identified 54 genome-wide significant loci, including genes linked to lens biology (Choquet, H. *et al*., 2021).

The prevalence of cataract varies by sex, geography, and ethnicity (Desai, N., & Copeland, R. A., 2013; World report on vision, 2019; Hashemi, H. *et al*., 2020; Fang, R. *et al*., 2022). Studies of cataract prevalence among Hispanics/Latinos (HL) suggest different prevalences among HL subgroups. A population-based study of Hispanic individuals aged 40 years or older found higher age-specific rates of significant cataract and cataract surgery among US Hispanic individuals than in African-American and European-ancestry individuals (Broman, A. T. *et al*., 2005). However, most GWAS on cataract have been performed in populations of European and Asian descent (Liao, J. *et al*., 2014; Ritchie, M. D., 2014; Yonova-Doing, E. *et al*., 2020; Hsu, C. C. *et al*., 2022). Consequently, the genetic susceptibility to cataract formation among HL remains unclear.

To better understand the genetic basis of cataracts in the HL population, we performed a GWAS of cataracts in more than 78,000 Puerto Rican research-consented participants of 23andMe. To maximize discovery power, we sequenced genomes of 500 Puerto Rican individuals to construct an imputation panel. We identified a significant association of large effect (OR=12.7, 95% CI=[6.5, 24.7]) at rs200560853, a rare coding variant in the ITGA6 gene. The risk allele is also associated with a 13.7 year earlier disease onset. We found that the risk variant was present at low frequencies in the Americas and in countries bordering the Mediterranean Sea, and absent in the rest of the world. Our results demonstrate the power of genetic studies in underrepresented and founder populations for improving our understanding of disease, and emphasize the need for their increased representation in genomics research.

## Results

### Identifying the Puerto Rican population and founder effect

We defined the Puerto Rican population in the 23andMe research cohort using a combination of self-reported grandparent birth-location and genetic relatedness based on identity-by-descent (IBD). First, we identified a set of 200 individuals who reported having 4 grandparents from Puerto Rico as reference individuals for the Puerto Rican population. Using the approach from Henn, B. M. *et al*. (2012), we computed IBD sharing between the reference individuals and used the results to define three features for each reference individual summarizing their genetic relatedness to the remaining reference individuals (Methods). We performed kernel-density estimation (KDE) using these features to learn a probability distribution over the reference set. To allow for error in self-reporting ancestry, we chose a probability threshold that excluded 10% of the reference individuals that were assigned the lowest probability by the KDE, as the minimum probability required for inclusion in the Puerto Rican population. We then applied the KDE to all research-consented 23andMe customers who shared any IBD with the 200 reference individuals, and identified 78,050 research participants as belonging to the same population, which we defined as being the Puerto Rican population for subsequent analyses.

To verify that the 23andMe Puerto Rican cohort showed a founder effect as has been previously reported, we used the IBDNe software (Browning, S. R., & Browning, B. L., 2015) to estimate recent effective population size of the Puerto Rican reference set (n=200) (Supplementary Figure 1). The analysis suggested a strong bottleneck in the Puerto Rican population approximately 8–12 generations ago, with the smallest effective population size (Ne) dating back approximately 10 generations ago (estimated Ne=1,470, 95% CI=[796, 2090]). These estimates are broadly consistent with previously reported estimates of the timing of the bottleneck, as well as their strength in Puerto Rican demographic history (Belbin, G. M. *et al*., 2017; Browning, S. R. *et al*., 2018; Mooney, J. A. *et al*., 2018).

### GWAS of cataracts

To discover genetic loci influencing cataract susceptibility in Puerto Ricans, we first undertook a GWAS of self-reported cataract status in the 23andMe Puerto Rican cohort. Participants included 3,060 cataract cases and 41,890 cataract-free controls, 80% of whom were over 30 years old and 56% were female (Supplementary Table 1). We identified three genome-wide significant loci (p-value < 5×10^−8^, see Figure 1a and Table 1), and the quantile-quantile plot indicated that type I error inflation was well controlled (genomic control factor = 0.93; Figure 1b). Since two out of the three lead variants were rare (minor allele frequency < 0.5%), which can affect the performance of the likelihood ratio test (LRT; Ma, C. *et al*., 2013), we also tested the three lead variants using the score test (Dey, R. *et al*., 2017). We found that only one of the three signals remained genome-wide significant when we applied the score test (Table 1). Therefore, we restricted our followup analysis to the single lead signal which is significant with both the LRT and score test.

**Figure 1a:**
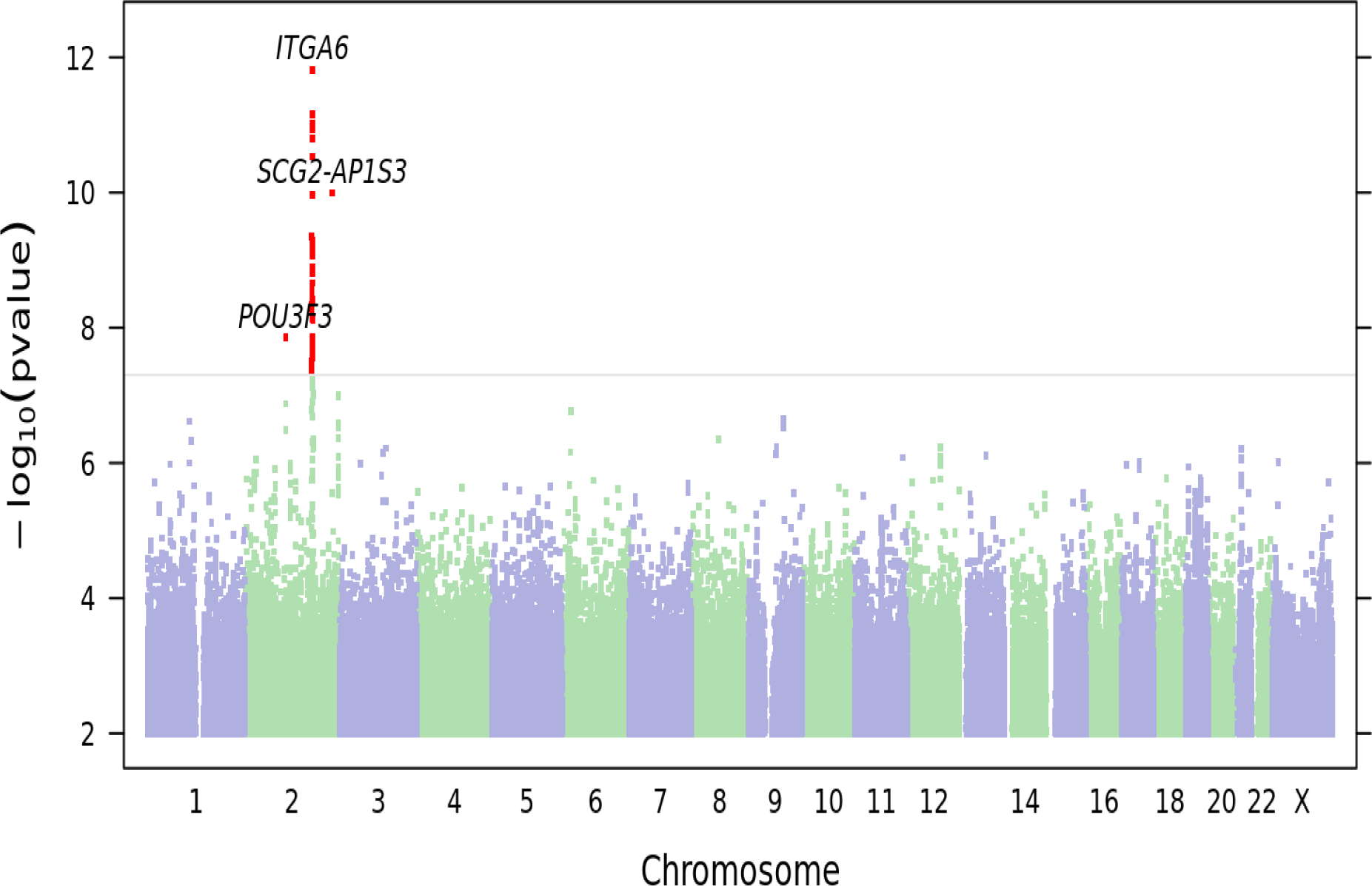
Manhattan plot showing results from the genome-wide association analysis of cataract. Three loci reach genome-wide significance (*P* ≤ 5e-8). Top-associated variants at each locus are labeled with their nearest gene context.

**Figure 1b:**
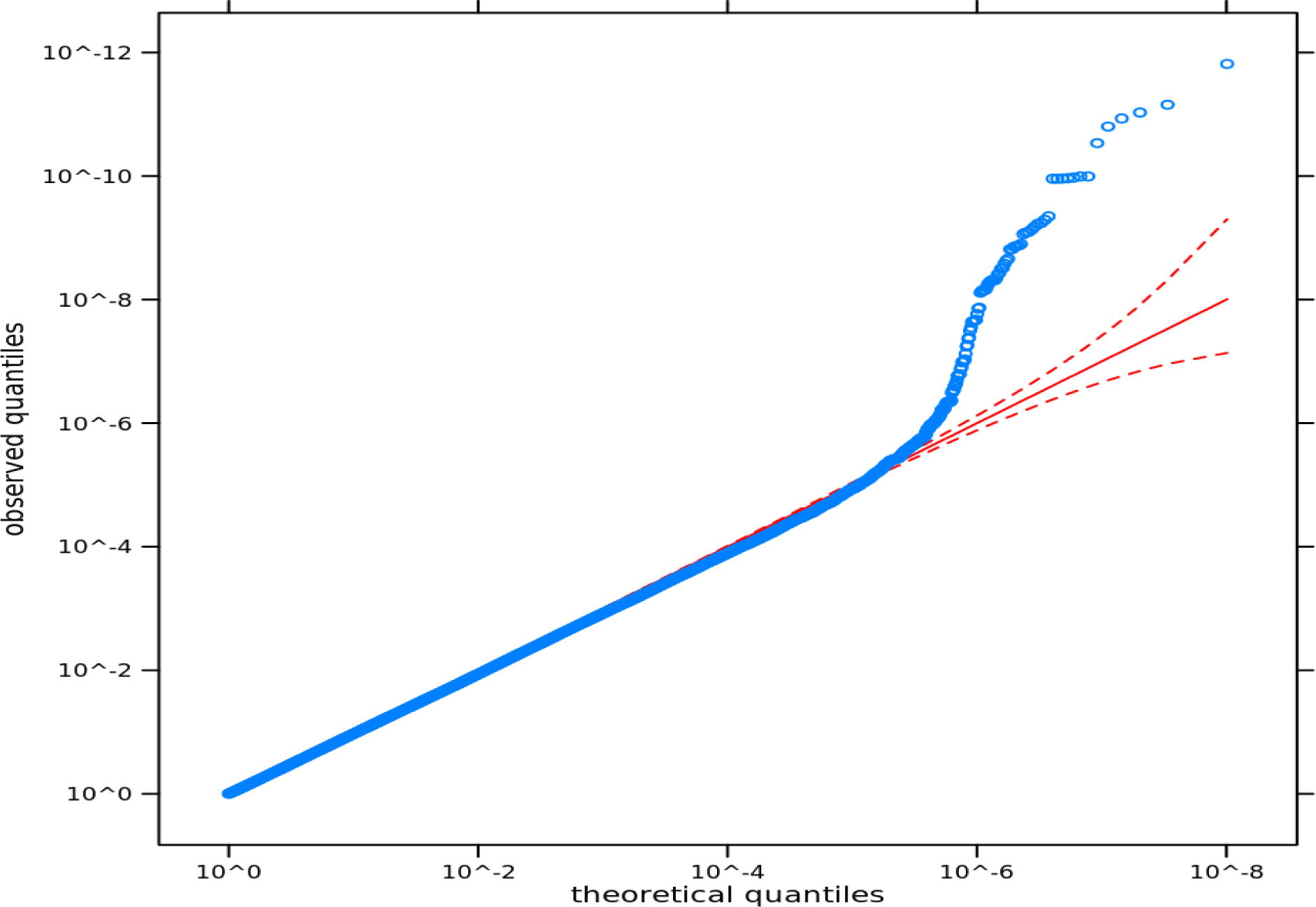
Q-Q plot of observed versus expected quantiles for the GWAS p-values, where the expected distribution of p-values is uniform under the null hypothesis, plotted on a log scale. A solid red line is shown with a slope of 1, and dashed red lines represent a 95% confidence envelope under the assumption that the test results are independent.

**Figure 2a:**
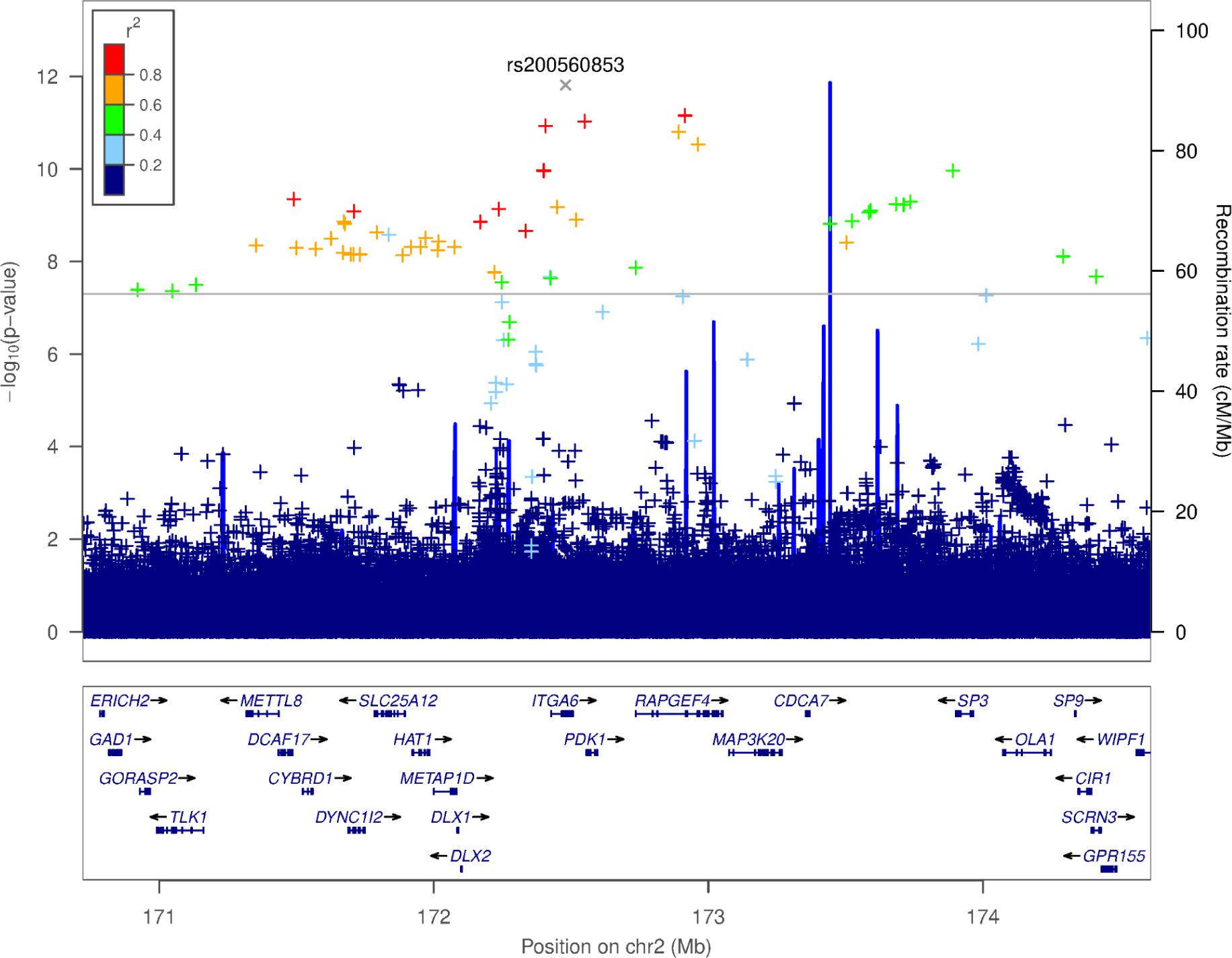
LocusZoom plot of the cataract GWAS test statistics versus position in the vicinity of the missense variant rs200560853. In the plot, a ‘+’ indicates an imputed non-coding variant, whereas a ‘x’ indicates an imputed protein-altering variant.

**Figure 2b:**
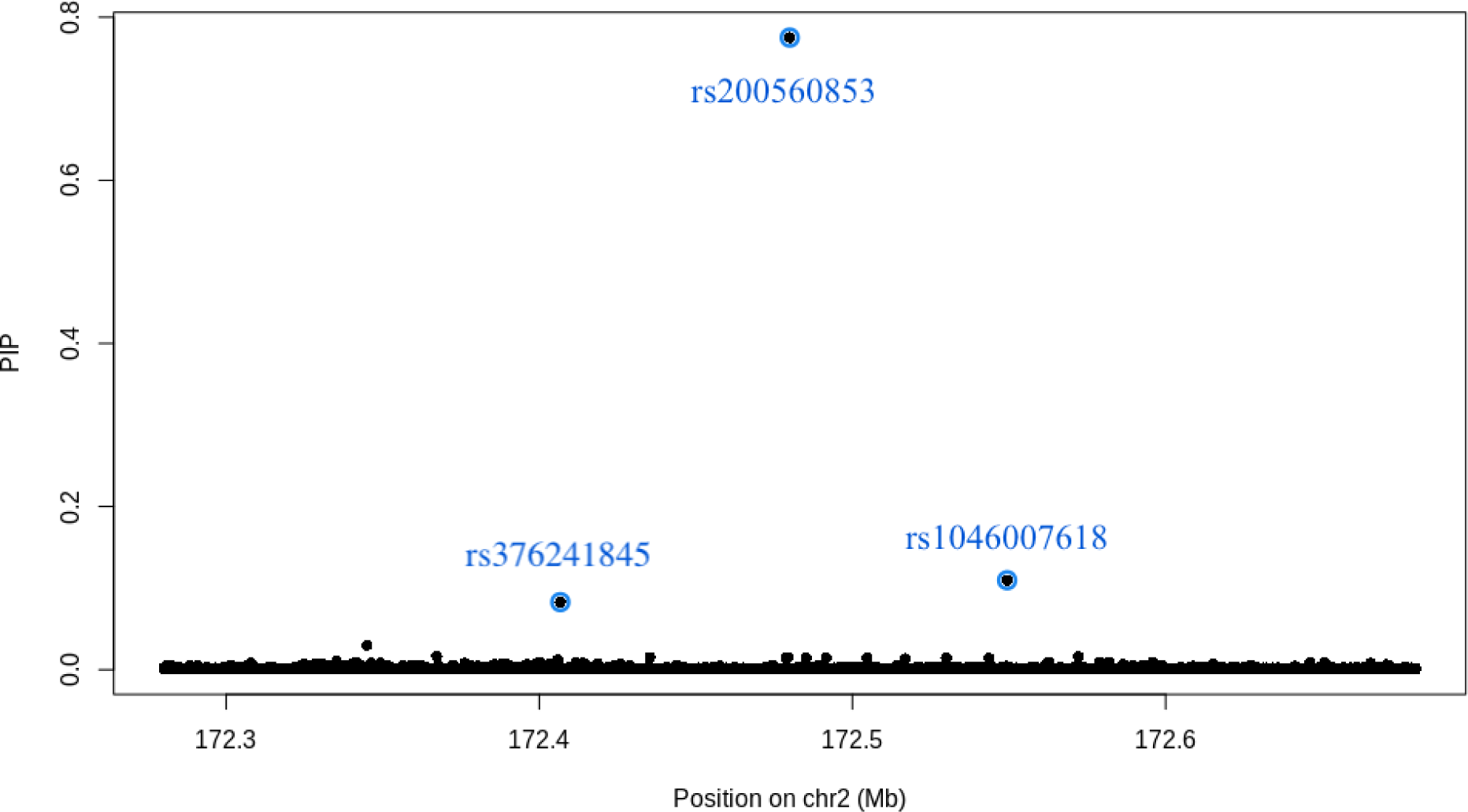
SuSiE PIP plot of posterior inclusion probability versus position in the ITGA6 locus. In the plot, the points highlighted in blue circles are variants selected by the SuSiE algorithm to form the 95% credible set.

**Figure 3a:**
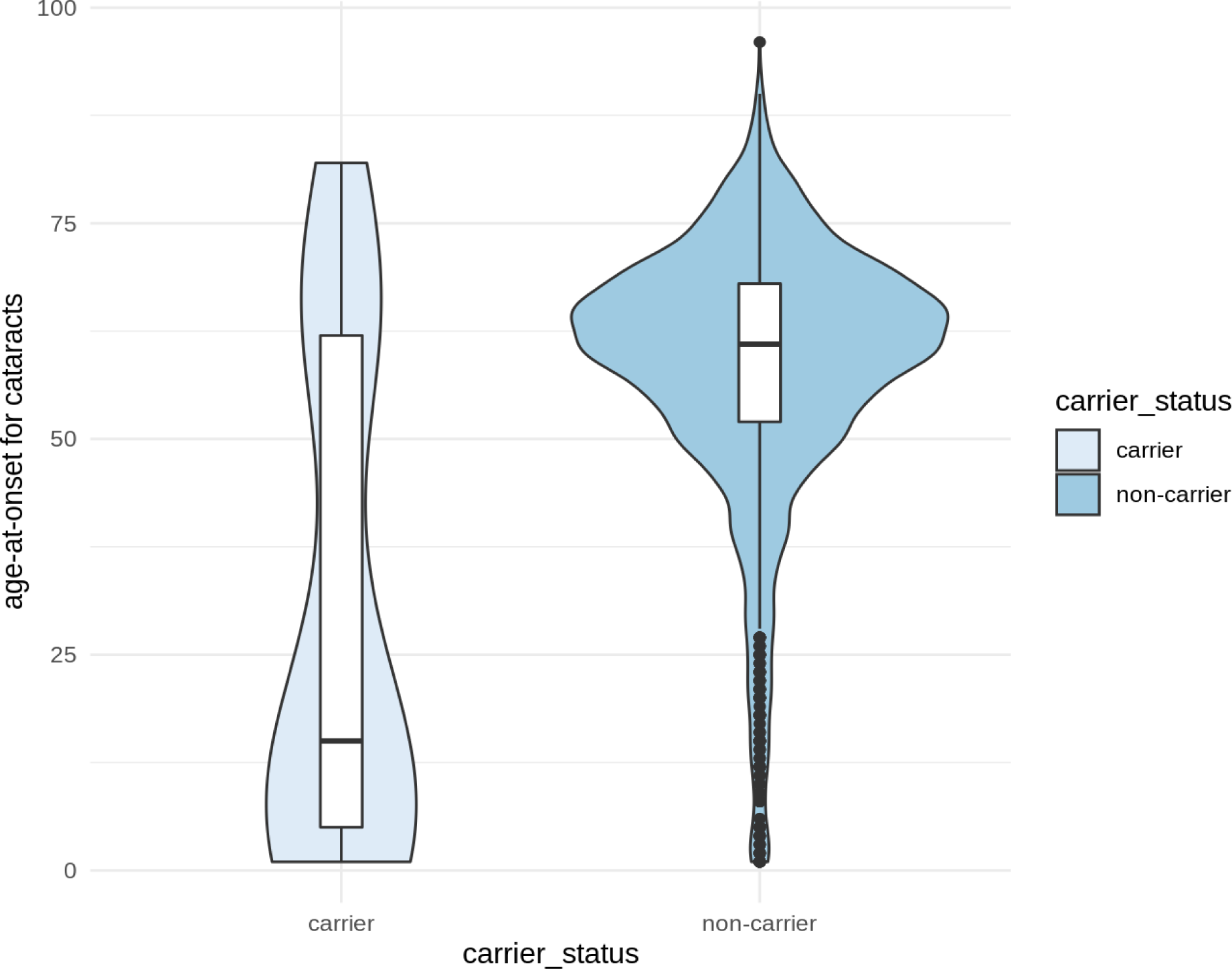
Violin plot of the age-at-onset data among the cataract cases. The left group represents 21 heterozygous carriers of the rs200560853 missense variant, the right group represents 2,332 non-carriers of the missense variant.

**Figure 3b:**
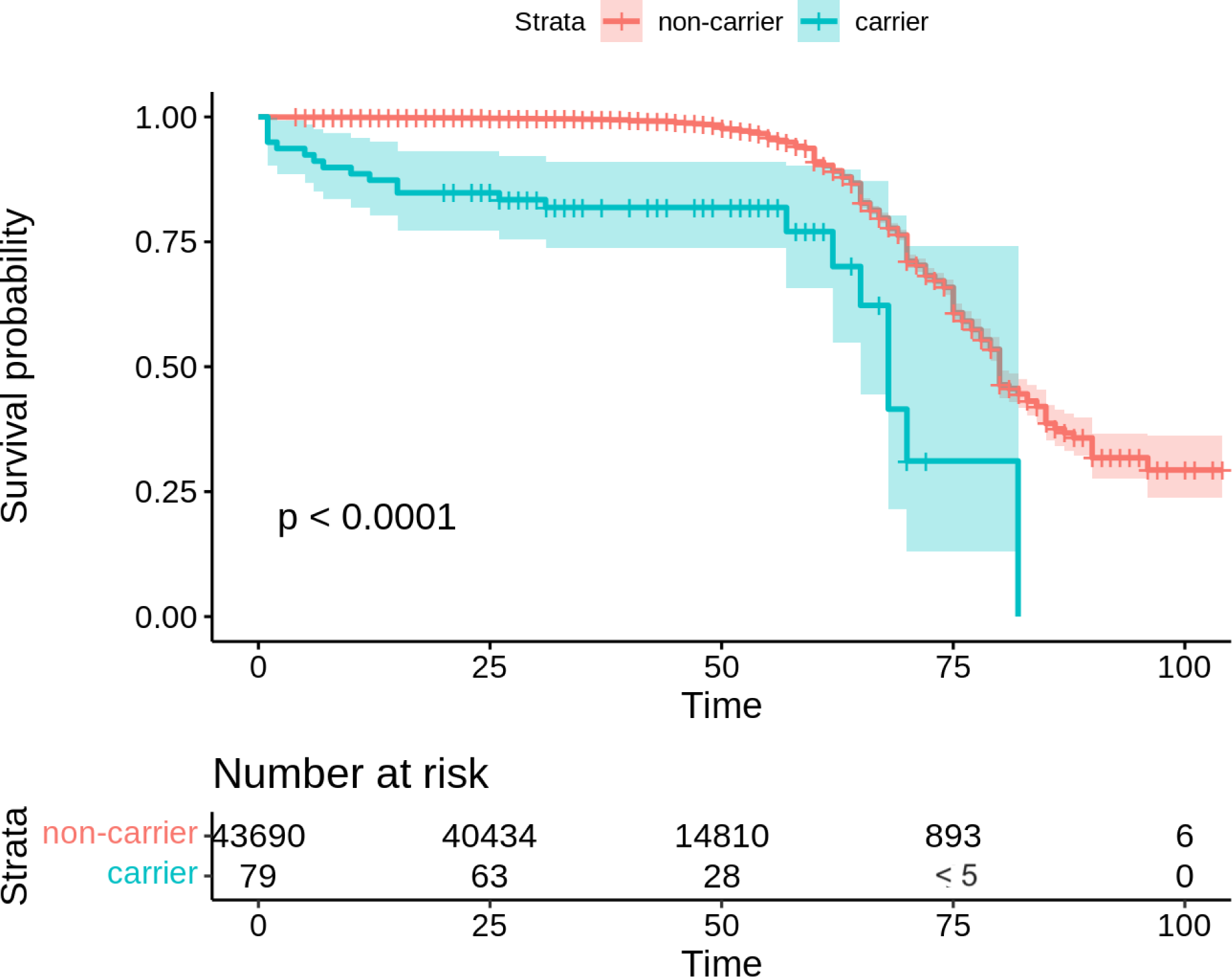
Kaplan-Meier curve for the time-to-cataract events. The green line represents heterozygous carriers of the rs200560853 missense variant, whereas the red line represents non-carriers of the missense variant.

**Table 1:** Genome-wide significant loci identified in the cataract GWAS (LRT: likelihood ratio test; OR: odds ratio, with respect to the effect allele; CI: confidence interval; EAF: effect allele frequency in Puerto Ricans)

| rsID | Cytoband | Position<br>(GRCh38) | Effect<br>Allele | Ref<br>Allele | Consequence | P-value<br>(LRT) | P-value<br>(score test) | OR | 95% CI | EAF |
| --- | --- | --- | --- | --- | --- | --- | --- | --- | --- | --- |
| rs200560853 | 2q31.1 | 172,479,990 | G | C | Missense & splice<br>variant within ITGA6 | $2.85 \times 10^{-12}$ | $4.01 \times 10^{-15}$ | 12.67 | [6.49, 24.77] | $8.89 \times 10^{-4}$ |
| rs145610699 | 2q36.1 | 223,706,185 | C | T | Intergenic between<br>SCG2 and-AP1S3 | $1.01 \times 10^{-10}$ | $5.28 \times 10^{-7}$ | 0.022 | $[2.9 \times 10^{-3}, 0.17]$ | $3.78 \times 10^{-3}$ |
| rs34791969 | 2q12.1 | 104,631,813 | A | G | Intergenic, upstream<br>of POU3F3 | $1.40 \times 10^{-8}$ | $1.28 \times 10^{-6}$ | 0.68 | [0.60, 0.78] | $8.07 \times 10^{-2}$ |

The variant with the strongest association with cataract was rs200560853 (NC_000002.12:g.172479990C>G, NG_008853.1:g.57405C>G), a missense and splice region variant in the ITGA6 gene on chr2q31.1 (LRT p-value=2.9×10^−12^; OR=12.7 [6.5, 24.7]; score test p-value=4.0×10^−15^; see Figure 2a). This signal appears to be novel as it has not been reported to be genome-wide significant in previous studies of cataract. Bayesian fine-mapping with SuSiE (Wang, G. *et al*., 2020) assigned the lead variant a high posterior inclusion probability (PIP) of being causal (PIP=0.775; see Figure 2b and Supplementary Table 2). The variant is well-imputed (imputation r^2^ = 0.99), despite being rare (MAF = 0.089%) in our Puerto Rican research cohort.

### Association with age of onset

We examined the association of rs200560853 with age-at-onset for cataract in a case-only analysis and a time-to-cataract survival analysis of the full cohort. Among the 2,353 cases who reported their age at cataract onset, we observed that the missense variant was significantly associated with an earlier onset of the disease (p-value=6.1×10^−11^, effect size = −13.7 years, 95% CI=[-17.8, −9.6]). Figure 3a depicts the distribution of age at cataract onset between the 21 heterozygous carriers and 2,332 non-carriers of the missense variant, demonstrating that the carrier group tends to have an earlier disease onset.

We also investigated the association of rs200560853 with time-to-cataract in the entire Puerto Rican cohort, with cataract cases who provided their age at cataract onset being classified as the events (n=2,353), and cataract controls being right-censored at their current age (n=41,416). Cox regression analysis confirmed that the missense variant was associated with a higher rate of cataract events (p-value=10^−7^, HR=4.3, 95% CI=[2.8,6.7]). Among the individuals analyzed in the Cox regression model, 79 were heterozygous carriers of the missense variant; 43,690 were non-carriers. The Kaplan-Meier analysis yielded a significant difference (Log-rank p-value < 0.0001) between the carrier vs non-carrier groups (Figure 3b).

### Effect of rs200560853 on the ITGA6 protein

According to the Human Protein Atlas, expression of gene ITGA6 has low tissue specificity and mRNA is detected in all tissues, including in the retina with an average (mean ± std) mRNA expression 7.3 ± 4.8 nTPM (Uhlén, M. *et al*., 2015). The rs200560853 C>G substitution alters the first nucleotide after the intron-exon junction (see Figure 4a) and the third base of codon TGC that spans exons ENSE00001073191 and ENSE00001073186 and encodes amino acid cysteine (Cys).

**Figure 4:**
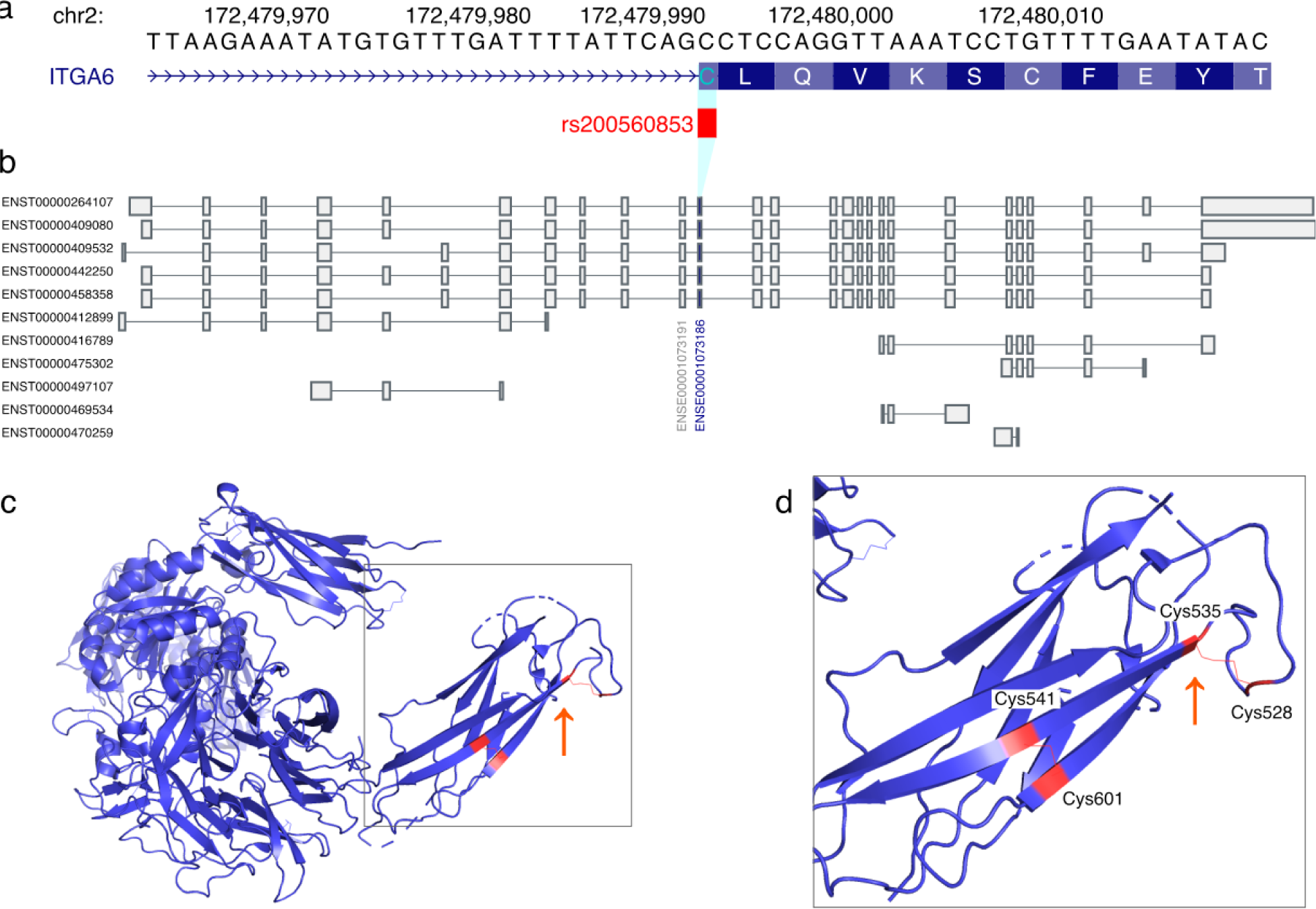
Position of variant rs200560853 (ITGA6 p.Cys535Trp) within a) exon ENSE00001073186, b) ITGA6 transcript models, and c) corresponding residue Cys535 in the protein structure of ITGA6 (PDB:7CEB). Inset d) shows the two disulfide bonds in the thigh domain, between Cys535 and Cys528, and between Cys541 and Cys601. Figures were downloaded from the a) UCSC Genome Browser and b) GTEx Portal, or c) d) generated by pymol, and subsequently modified.

Based on information from the gnomAD resource (v3.1.2), SpliceAI acceptor loss ΔS for the rs200560853 variant is 0.02 (Jaganathan, K. *et al*., 2019), suggesting that splicing is not likely to be a relevant mechanism of action for how rs200560853 influences early-onset cataract. Due to the population specificity and low MAF of rs200560853, we were unable to quantify any potential differences in numbers of reads spanning the intron-exon junction in carriers of C and G alleles, as it was not possible to detect this variant in data from the GTEx Consortium (GTEx Consortium., 2020).

Missense variant rs200560853 C>G changes Cys into tryptophan (Trp) in all protein coding transcripts that contain exons ENSE00001073191 and ENSE00001073186 (see Figure 4b and Supplementary Table 4). According to UniProt, in the main protein isoform of P23229 (corresponding to transcript ENST00000442250) Cys535 is located in the ectodomain of ITGA6 and is predicted to form a disulfide bond with nearby Cys528; this is further substantiated by the crystal structure of the ITGA6 fragment deposited in the Protein Data Bank (PDB accession codes 7CEB, 7CEC; (Arimori, T. *et al*., 2021)) which shows a disulfide bond between residues Cys528 and Cys535 (see Figure 4d). Disulphide bonds in the ectodomain of integrins carry out either structure-stabilizing or regulatory functions by acting as reversible thiol switches (Lorenzen, I., Eble, J. A., & Hanschmann, E. M., 2021), and disruption of the Cys528-Cys535 disulfide bond by the amino acid change p.Cys535Trp may have an impact on the structure of the extracellular thigh domain or on the ability of ITGA6 to switch between bent, extended closed and extended open confirmations, thus leading to obstruction of ligand binding.

### Geographical distribution and putative origin of the ITGA6 missense variant

The ITGA6 missense variant is rare in public databases. In gnomAD (v 3.12), it is present in the Latino/Admixed American population with a MAF of 0.01965% (3 allele count among 15,268 measured alleles) and is unobserved in any of the non-Latin populations, with an allele count of 0 among 136,862 alleles (Supplementary Table 3).

To identify other populations that may carry the ITGA6 risk variant, we examined its worldwide geographical distribution through imputation of a larger subset of the 23andMe cohort. To reliably assign allele frequencies to geographical divisions, we considered 23andMe consented research participants who were genotyped on the v5 (GSA-based) array and answered survey questions reporting that their four grandparents were all born within one country/geographic division (excluding the US and Canada, henceforth referred to as the 4GP cohort, see Methods). This cohort comprises 1,091,730 individuals from 228 countries/geographic divisions across the world. The allele frequency for rs200560853 was non-zero in 20 out of 228 worldwide geographic divisions (Table 2). Globally, we found that the frequency of rs200560853 was greater than 0.1% in only two geographic divisions: Puerto Rico (0.105%, 95% CI = [0.074%, 0.141%]) and Morocco (0.121%, 95% CI = [0.008%, 0.271%]) (Methods), with a wide confidence for the estimate in Morocco due to its small 4GP cohort size.

**Table 2:**
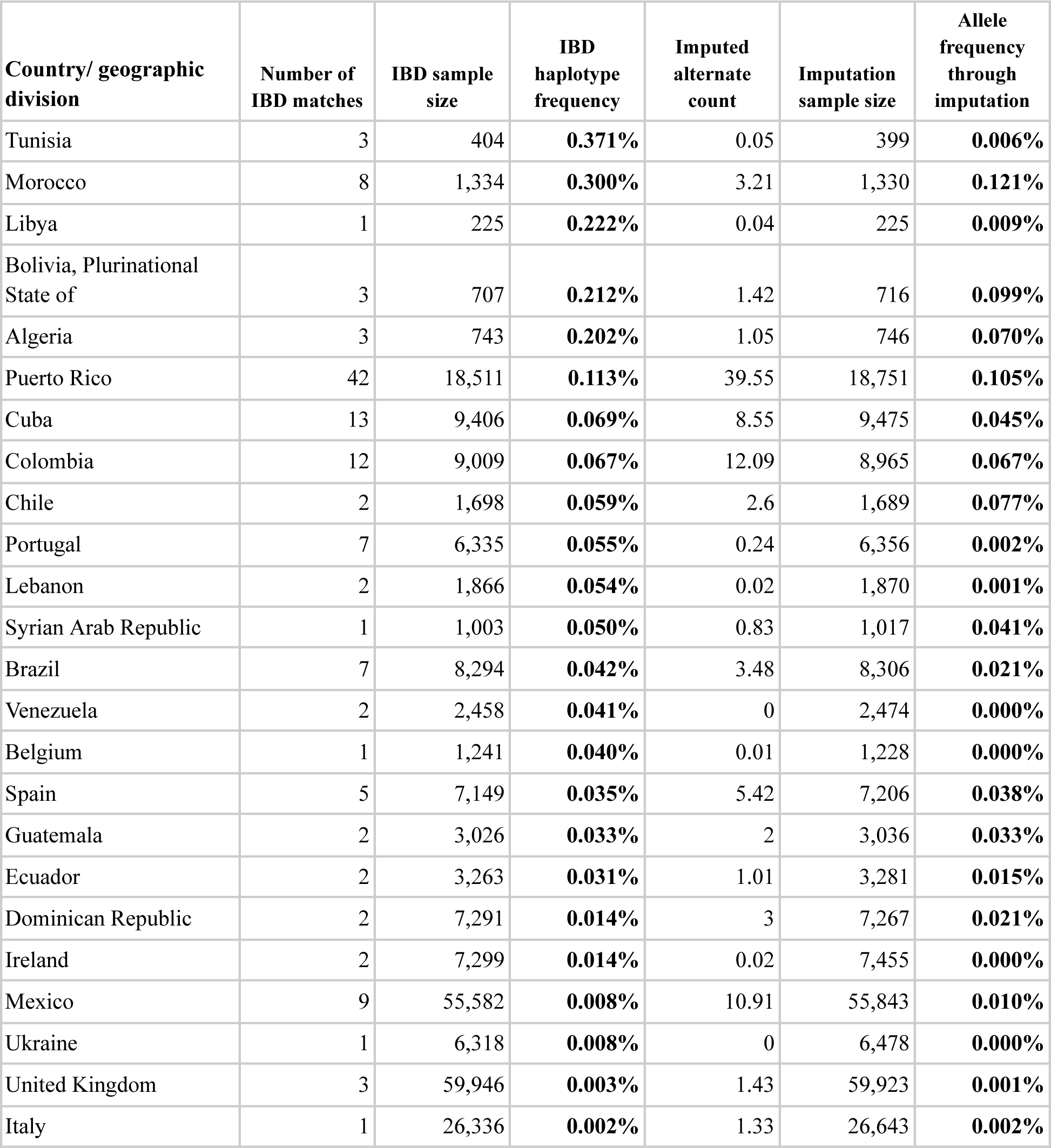
Inference of allele frequency using IBD with carrier individual haplotypes across the 300-SNP Ancestry Composition window, and through imputation of the missense allele, against a reference panel of 23andMe research participants with all four grandparents born in a single country/geographic division (exempting US and Canada).

| **Country/ geographic division** | **Number of IBD matches** | **IBD sample size** | **IBD haplotype frequency** | **Imputed alternate count** | **Imputation sample size** | **Allele frequency through imputation** |
| --- | --- | --- | --- | --- | --- | --- |
| Tunisia | 3 | 404 | **0.371%** | 0.05 | 399 | **0.006%** |
| Morocco | 8 | 1,334 | **0.300%** | 3.21 | 1,330 | **0.121%** |
| Libya | 1 | 225 | **0.222%** | 0.04 | 225 | **0.009%** |
| Bolivia, Plurinational State of | 3 | 707 | **0.212%** | 1.42 | 716 | **0.099%** |
| Algeria | 3 | 743 | **0.202%** | 1.05 | 746 | **0.070%** |
| Puerto Rico | 42 | 18,511 | **0.113%** | 39.55 | 18,751 | **0.105%** |
| Cuba | 13 | 9,406 | **0.069%** | 8.55 | 9,475 | **0.045%** |
| Colombia | 12 | 9,009 | **0.067%** | 12.09 | 8,965 | **0.067%** |
| Chile | 2 | 1,698 | **0.059%** | 2.6 | 1,689 | **0.077%** |
| Portugal | 7 | 6,335 | **0.055%** | 0.24 | 6,356 | **0.002%** |
| Lebanon | 2 | 1,866 | **0.054%** | 0.02 | 1,870 | **0.001%** |
| Syrian Arab Republic | 1 | 1,003 | **0.050%** | 0.83 | 1,017 | **0.041%** |
| Brazil | 7 | 8,294 | **0.042%** | 3.48 | 8,306 | **0.021%** |
| Venezuela | 2 | 2,458 | **0.041%** | 0 | 2,474 | **0.000%** |
| Belgium | 1 | 1,241 | **0.040%** | 0.01 | 1,228 | **0.000%** |
| Spain | 5 | 7,149 | **0.035%** | 5.42 | 7,206 | **0.038%** |
| Guatemala | 2 | 3,026 | **0.033%** | 2 | 3,036 | **0.033%** |
| Ecuador | 2 | 3,263 | **0.031%** | 1.01 | 3,281 | **0.015%** |
| Dominican Republic | 2 | 7,291 | **0.014%** | 3 | 7,267 | **0.021%** |
| Ireland | 2 | 7,299 | **0.014%** | 0.02 | 7,455 | **0.000%** |
| Mexico | 9 | 55,582 | **0.008%** | 10.91 | 55,843 | **0.010%** |
| Ukraine | 1 | 6,318 | **0.008%** | 0 | 6,478 | **0.000%** |
| United Kingdom | 3 | 59,946 | **0.003%** | 1.43 | 59,923 | **0.001%** |
| Italy | 1 | 26,336 | **0.002%** | 1.33 | 26,643 | **0.002%** |

To investigate the putative ancestral origin of rs200560853, we examined Ancestry Composition results computed from phased genotype array data of risk allele carriers. We estimated local ancestry across the ITGA6 locus in individuals carrying the risk allele (Figure 5). We also examined local ancestry in a similar sample of individuals, who reported that their four grandparents were born in Puerto Rico, but who were not inferred to be carriers for the risk allele. We observe an increase in estimated North African ancestry among carriers for the window containing the risk allele, which is not seen in the random sample of non-carrier Puerto Rican individuals. This increase is seen in individuals genotyped on both platforms (Supplementary Figure 2), and is far above the genome-wide average, suggesting that the background haplotype on which the risk allele is located may be found in present-day individuals of Algerian, Libyan, Moroccan, Mozabite, and Tunisian descent, supporting a North African origin.

**Figure 5:**
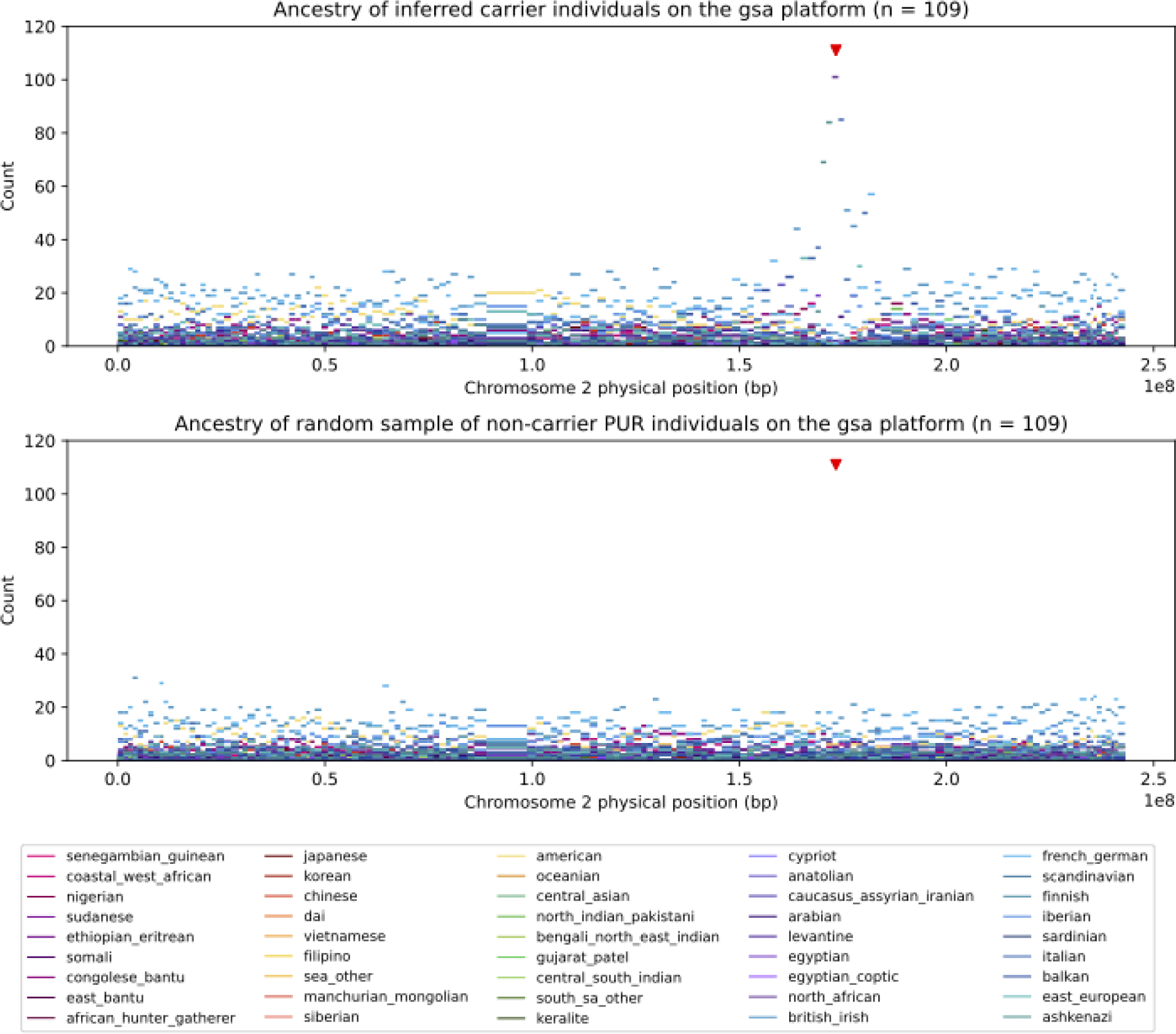
Ancestry of inferred carrier individuals’ haplotypes genotyped on the GSA-based array (top) and a random subsample of Puerto Rican non-carrier individuals (bottom) along chromosome 2. Red triangle indicates the location of the rs200560853 missense variant. The x-axis corresponds to the physical position of each 300-SNP Ancestry Composition window, and the y-axis shows the number of windows assigned to be of each ancestry, designated by color. The carrier individuals show a clear peak of North African ancestry at the window overlapping the locus of interest, while non-carrier individuals do not.

As an additional line of evidence supporting a North African origin of the missense variant, we computed Identical-by-Descent (IBD) segments using phasedIBD on the carriers’ genotype array data (Freyman, W.A. *et al*., 2021) against the 4GP cohort. We find that the carrier individuals’ haplotypes across the window of interest most frequently match 4GP individuals who reported their grandparents to have been born either in the Americas or Caribbean or countries bordering the Mediterranean Sea (Table 2). The highest frequencies inferred for the background haplotype across this window, outside of the Americas, are in individuals with four grandparents from Tunisia, Morocco, Libya, and Algeria. Overall, the frequency of haplotype matches showed a moderate correlation with the number of IBD matches to the Puerto Rican carriers (Spearman correlation coefficient r^2^=0.564; Pearson correlation coefficient r^2^=0.451). The haplotype frequencies are also significantly higher for most countries than the estimated allele frequencies from imputation, suggesting that the missense allele may be polymorphic on an ancestral background haplotype in these populations.

## Discussion

In this study, we carried out a genome-wide association analysis of cataract in 23andMe research participants of Puerto Rican ancestry, identified via self-reported grandparent birth-location and genetic relatedness. We discovered a novel association that confers a 12-fold increased risk of cataract and fine-mapped it to a missense variant in the ITGA6 gene. We also found that the missense variant was significantly associated with earlier disease onset among cataract cases, as well as an increased rate of cataract events in the case-control cohort. These findings suggest that the ITGA6 missense variant may be a novel genetic risk factor for cataract in Puerto Ricans.

The ITGA6 rs200560853 missense variant is rare (MAF = 0.089%) in our Puerto Rican cohort. By sequencing only a few hundred individuals of Puerto Rican ancestry, we were able to impute it to a high degree of accuracy in the 23andMe cohort. This, combined with the large sample size of the Puerto Rican GWAS, allowed the discovery of the association of the missense variant with cataract susceptibility and age-of-onset. The low global frequency of this variant could explain why this association has not been previously reported.

Patterns of genetic diversity in Puerto Ricans indicate a strong founder effect and population bottleneck in the recent past (Ayala, C. J. *et al*., 2009; Belbin, G. M. *et al*., 2017; Browning, S. R. *et al*., 2018; Mooney, J. A. *et al*., 2018). Founder populations exhibit an excess of private and low frequency variants above that expected under Wright-Fisher models (Coventry, A. *et al*., 2010; Keinan, A., & Clark, A. G., 2012; Tennessen, J. A. *et al*., 2012; Casals, F. *et al*., 2013). Previous studies in Puerto Ricans have identified founder mutations that affect disease risk (Anikster, Y. *et al*., 2001; Gonzaga-Jauregui, C. *et al*., 2015). Our finding of a large-effect globally rare deleterious variant associated with cataract is consistent with a founder effect in Puerto Rico, and we find evidence for this in our data (Supplementary Figure 1). Those examples and our findings highlight the importance of genomic studies in underrepresented populations, and particularly in founder populations.

Using the 23andMe cohort, we were able to infer the worldwide geographic distribution of the missense variant through imputation and IBD analyses. Both analyses found that the variant was present at low frequencies in the Americas, the Caribbean or in countries bordering the Mediterranean Sea. In addition, local ancestry in a window around the missense variant showed a large increase in North African ancestry among carriers, suggesting a likely North African origin for the missense variant. Genome-wide, North African ancestry among the Puerto Rican carriers is generally low (1.5% on average). However, migration from the Canary Islands into Puerto Rico is well documented (Parsons, J. J., 1983), and previous mtDNA studies suggest that the Canary Islands are likely a major contributing source to the North African and West Eurasian ancestry present in Puerto Ricans (Díaz-Zabala, H. J. *et al*., 2017). The aboriginal inhabitants of the Canary Islands, Guanches, are shown to have North African ancestry (Fregel, R. *et al*., 2009, Rodríguez-Varela, R. *et al*., 2017), possibly from a Berber or Moorish source. However, North African ancestry in the carrier individuals could have many other possible sources, including through Spain, as the Iberian peninsula is shown to have higher levels of North African admixture than other European populations (typically estimated at about 4% - 10% admixture) (González, A. M. *et al*., 2003; Moorjani, P. *et al*., 2011; Botigué, L. R. *et al*., 2013). Prior haplogroup studies suggest that Moorish invasions have left a Jewish and Arab/Berber genetic influence in southern Iberia (Adams, S. M. *et al*., 2008). Our results are therefore only suggestive and not conclusive of a North African origin for the missense variant.

The novel association we found was fine-mapped to the ITGA6 gene, an integrin alpha chain in the laminin receptor subfamily that associates with ITGB1 or ITGB4 to form ɑ6β1 or ɑ6β4 heterodimeric receptors. Within the eye lens, ITGA6 is primarily found in lens fiber cells while ITGB1 is common to all lens membrane surfaces. (Menko, A. S., & Philip, N. J., 1995). Previous studies have shown biological roles for ɑ6β1 integrin in maintaining lens homeostasis, transparency, as well as lens cell survival (Menko, A. S., & Andley, U. P., 2010; Liu, J. *et al*., 2020). ɑ6β1 integrin plays a key role in the microcirculation of glutathione and glucose across mouse lens cortical fibers (Liu, J. *et al*., 2020), two circulating components shown to protect lens fibers against oxidative stress (Braakhuis, A. J. *et al*., 2019). Moreover, ɑ6β1 has been shown to associate with the heat shock protein alpha A-crystallin in lens fiber cells *in vivo*, promoting lens fiber cell survival during differentiation (Menko, A. S., & Andley, U. P., 2010). Together, these studies suggest a possible explanation for how ITGA6 variants may increase cataract risk through multiple biological mechanisms. Confirmatory functional studies *in vitro* or in animal models are critical in order to characterize the molecular mechanisms underpinning the novel association that we identified, and these could help unravel how the missense variant contributes to the genetic predisposition to cataract. Variants in ITGA6 are reported to cause junctional epidermolysis bullosa-6 with pyloric atresia, an autosomal recessive blistering disease of skin and mucous membranes (Ruzzi, L. *et al*., 1997; Masunaga, T. *et al*., 2017). Ocular complications are commonly reported in epidermolysis bullosa subtypes, however, data on association with cataract is limited due to small sample sizes (Tong, L. *et al*., 1999; Fine, J. D. *et al*., 2004). Further studies are needed to evaluate a possible association between epidermolysis bullosa and cataract.

There are several limitations to our study. First, our study relied on self-reported cataract status and age-of-onset information. Though previous studies have demonstrated genetic correlation of 0.8-0.9 between self-reported and hospital record cataract phenotypes (DeBoever, C. *et al*., 2020), it is unclear how bias in self-reporting may have affected our results. Second, due to the low frequency of the lead variant, we were unable to find a replication cohort of sufficiently large size. External replication would help reduce concerns about phenotypic ascertainment/bias as well as the robustness of our association signal. Third, although the missense variant is associated with a more than twelve-fold increased risk of cataract, more than 65% of risk allele carriers remained free of disease at the time their survey responses were collected. Prospective studies of genetically at-risk individuals are needed to inform penetrance. Fifth, additional population genetic studies are needed to further clarify the evolutionary origin of the missense variant or the potential role of drift in determining its frequency.

Cataracts result in visual impairment, which is associated with challenges with activities of daily living, social activities, mobility, cognitive ability, longer nursing home stays, and falls (Desai, N., & Copeland, R. A., 2013). While cataracts are routinely treated with surgery, language and financial barriers are known to impede access to cataract surgery in

Hispanic/Latino populations (Broman, A. T. *et al*., 2005). Smoking, alcohol consumption, UV exposure, and nutrition are some of the modifiable non-genetic risk factors for cataract. The large effect of the ITGA6 missense variant, with a 12-fold increased cataract risk and 13 years earlier age-of-onset, along with its increased frequency in Puerto Rico and the Americas raises the possibility that screening for this variant in populations at risk may allow earlier detection of cataract. Lifestyle interventions targeting non-genetic cataract risk factors could then be targeted to individuals carrying the risk allele to reduce overall cataract risk or delay onset.

## Methods

### Research participants

Eligible research participants were selected from the 23andMe customer base who provided informed consent and volunteered to participate in the research online, under a protocol approved by the external AAHRPP-accredited IRB, Ethical & Independent (E&I) Review Services. As of 2022, E&I Review Services is part of Salus IRB (https://www.versiticlinicaltrials.org/salusirb). Inclusion criteria of the research participants was based on their consent status at the time when the data analysis was initiated.

### DNA Extraction and Genotyping

DNA extraction and genotyping of research participants were performed on saliva samples by Laboratory Corporation of America, the CLIA-certified and CAP-accredited clinical laboratories. Samples were genotyped on one of three genotyping platforms (which we refer to as v1, v2, v3, v4, and v5). The v1 and v2 platforms were based on the Illumina HumanHap550+ BeadChip, with a total of about 560,000 SNPs, including about 25,000 custom SNPs selected by 23andMe. The v3 platform was based on the Illumina OmniExpress+ BeadChip, with a total of about 950,000 SNPs. The v4 platform was a fully customized array, with a total of about 570,000 SNPs for the additional coverage of lower frequency coding variants. The v5 platform is in current use, which is an Illumina Infinium Global Screening Array with about 640,000 SNPs in total, including around 50,000 custom SNPs selected by 23andMe. When genotyping the saliva samples, those that failed to reach the 98.5% call rate were re-analyzed. For any customers whose DNA extraction and genotyping procedure failed repeatedly, they were re-contacted by 23andMe customer service to provide additional saliva samples.

### Founder event in the Puerto Ricans

We identified 200 individuals who self-reported having 4 grandparents from Puerto Rico. Using genotype array data for these individuals, we generated identity-by-descent (IBD) segments for all pairs using the method described in Henn, B. M. *et al*., 2012. We used the IBDNe program (Browning, S. R., & Browning, B. L., 2015) with the bootstrapping samples parameter (nboots) set to 50 to infer recent effective size from IBD segments in the 200 Puerto Rican samples.

### Puerto Rican population definition

To identify candidate individuals for inclusion in the Puerto Rican population definition, we first estimated a distribution over 200 individuals who self-reported having 4 grandparents from Puerto Rico (“reference individuals”) and then identified all other individuals who belonged to that distribution.

For each individual in the reference set, we defined three features based on their IBD sharing with the remaining reference individuals: 1. number of reference individuals with whom they shared any IBD; 2. median total genome-wide IBD with a reference individual; and 3. the median length of the longest IBD segment with a reference individual. Using these features for the 200 reference individuals, we performed kernel-density estimation using the *gaussian_kde* function in the *scipy* package (Virtanen, P. *et al*., 2020). We then evaluated the estimated probability density function on the 200 reference individuals to compute a probability of belonging to the estimated distribution. As a threshold for inclusion of reference individuals in the distribution, we chose the 10th percentile, to allow for error in self-reporting ancestry and to include 90% of the reference individuals in the distribution.

To identify additional individuals who belong to the estimated distribution, we considered all individuals (excluding the reference individuals) who had any IBD with the 200 reference individuals chosen above. We computed the 3 features described above for each candidate individual, and evaluated the estimated probability density function for those feature values. If the estimated probability was larger than the threshold value, we classified this candidate individual as belonging to the Puerto Rican population. With this approach, 78,050 research-consented participants from the 23andMe customer base were identified as Puerto Ricans.

### Sequencing Puerto Rican genomes for an imputation panel

We selected 500 samples, chosen at random from the set of predicted Puerto Rican individuals, for whole-genome sequencing. The Puerto Rican individuals were sequenced to an average depth of 23x. All sequenced customers consented to research participation. These were combined with high coverage sequencing from 1000 Genomes (N=2,504) (Byrska-Bishop, M. *et al*., 2022), HGDP/SGDP (N=969) (Bergström, A. *et al*., 2020) and GTEx-v8 (N=838) (GTEx Consortium., 2020) to yield an ethnically diverse imputation panel containing 13,599 individuals. Sequencing reads were aligned to GRCh38 (Schneider, V. A. *et al*., 2017) with BWA-MEM (0.7.15) (Li, H., 2013) and post-processed as described in the functional equivalence pipeline (Regier, A. A. *et al*., 2018). Variant calling was performed with DeepVariant (0.8.0) (Poplin, R. *et al*., 2018) and GLnexus (1.2.7) (Lin, M. F. *et al*., 2018; Regier, A. A. *et al*., 2018). Multiallelic variants were split and normalized with the “bcftools norm -m -any -f ref.fa” command. Genotypes with GQ<20 were set to missing and subsequently any variant with any of >20% missingness, Inbreeding coefficient < −0.3 or allele count ≤1 was removed. Finally, the resulting multi-sample VCF was phased using SHAPEIT4 (Delaneau, O. *et al*., 2019).

### Genotype imputation

We phased research participants on the v1/v2 chip, v3 chip and v4 chip as separate cohorts using SHAPEIT4. The v5 cohort was too large to phase jointly, so an initial batch of one million research participants was phased using SHAPEIT4. This was then used as a reference panel to phase subsequent smaller batches of individuals (with new batches arriving on an approximately weekly cadence). These pre-phased data were then imputed using the previously described reference panel and Beagle5 (Browning, B. L. *et al*., 2018). The final imputation panel contained 117,772,876 variants.

### Phenotype definitions for GWAS

We collected self-reported cataract diagnosis history from 23andMe research participants using a web-based survey, and used information derived from the survey to define phenotypes for the GWAS. Specifically, participants were asked to respond to the question “Have you ever been diagnosed with, or treated for, cataracts?”, with possible responses “Yes / No / I’m not sure”. Of those who responded “Yes”, we further asked the question “How old were you when you were first diagnosed with cataracts? Your best guess is fine.”, to which participants could provide their age of diagnosis for cataracts.

We defined the binary cataract phenotype with cases as those who responded “Yes” to the cataract diagnosis or treatment survey question, and controls as those who answered “No” to that question. We defined the quantitative age-at-onset for cataract phenotype using the age values collected from the age at first diagnosis with cataract survey questions. We also defined the time-to-cataract survival phenotype using the age at first diagnosis with cataract as the event time, and we right-censored those who answered “No” to the cataract diagnosis or treatment survey question using their current age.

### GWAS modeling

We chose a maximal set of unrelated individuals when running the GWAS, such that no two individuals shared more than 700 cM of identity-by-descent (IBD), including regions where the two individuals share either one or both genomic segments IBD. If a cataract case and a cataract control were identified as IBD with each other, we preferentially kept the case in the sample and discarded the control. This resulted in approximately 19.8% of the sample being excluded from the cataract GWAS.

To account for population structure within the sample, we performed principal component analysis (PCA) using 63,470 high quality genotyped variants that are present in all five genotyping platforms on 78,394 samples.

For the cataract case-control comparison, we tested for the association using logistic regression; for the age-at-onset and time-to-cataract data, we ran linear and Cox regression for the association testing, respectively. We assumed additive allelic effects of the genotyped and imputed data in all regression models. Tests for the autosomes were based on the genotyped allele counts for the genotyped SNPs, and imputed allele dosages for the imputed SNPs. Tests for the X chromosome were computed similarly, with men coded as if they were homozygous diploid for the observed allele. In all regression models, we included covariates for age, sex, the top five principal components (PCs) to account for residual population structure, and dummy variables for genotype platforms to account for genotype batch effects.

We used the likelihood ratio test to compute the association test *P* values. When choosing between the genotyped and imputed results to combine the GWAS summary statistics, we favored the imputed result, unless the imputed SNP was unavailable or failed quality control (QC). For genotyped variants, our QC criteria removed v1/v2 variants (due to small sample size) and those that failed the transmission disequilibrium test in trios with *P* < 10^−20^, or failed the Hardy–Weinberg equilibrium (HWE) test in individuals of Puerto Rican ancestry with *P* < 10^−20^, or failed the batch effect test with *P* < 10^−50^ for the analysis of variance (ANOVA) *F*-test across batches, or had a genotyping call rate less than 90%. For imputed variants, we removed variants with low imputation quality (with the averaged *r*^2^ across batches < 0.5, or the minimum *r*^2^ < 0.3), or with evidence indicating batch effects such that ANOVA *P* < 10^−50^.

For each GWAS, we identified loci with genome-wide significant associations. We first gathered all SNPs with *P* < 10^−5^ within the vicinity of a genome-wide significant association to define the region boundaries, next we grouped these regions into loci so that two adjacent loci were separated by at least 250 kilobases (kb). We chose the SNP with the smallest *P* value in a locus as the lead SNP.

### Statistical fine-mapping

To identify genetic variants which causally affect cataract in the ITGA6 locus, we performed statistical fine-mapping with the SuSiE approach (Wang, G. *et al*., 2020), which accounts for correlations between SNPs due to high LD when selecting SNPs for the credible set construction.

We constructed an in-sample LD panel from the cataract GWAS samples and computed the LD matrix using plink2 (Chang, C. C. *et al*., 2015). We used the default parameter settings in SuSiE, which allows up to ten independent signals in the locus.

### Ancestry analysis

To investigate a putative ancestral origin of rs200560853, we examined Ancestry Composition estimates on chromosome 2, computed for the carriers’ genotype array data. The Ancestry Composition algorithm has been described previously (Durand, E. Y., 2021). Briefly, Ancestry Composition infers local ancestry assignments for 300-SNP windows across the genome via comparison with a large reference dataset using a Support Vector Machine (SVM) approach. Since Ancestry Composition (AC) estimates local ancestry for each statistically- (or parent-) phased chromosome, to determine the inferred ancestry of the haplotype carrying the rs200560853 risk variant, we first mapped the haplotype comprising the imputed sites from position (GRCh38) 172,280,147 to 172,679,872 onto the available array sites. We required an exact haplotype match across 99 sites (v5 GSA-based array) or 158 sites (v1-v4 versions) to identify the corresponding array-based phased haplotype. We then summed the results on the chromosome 2 haplotype from Ancestry Composition’s first stage, where a string-kernel support-vector-machines classifier assigns provisional ancestry labels to short genomic segments, across all haplotypes inferred to carry the rs200560853 risk allele. We used the first stage only, as each window is treated independently from neighboring windows, and provides the most point-like estimate of ancestry for the Ancestry Composition 300-SNP window which spans the locus of interest. Data for the first stage from AC version 5.5.2 were utilized. Since Ancestry Composition windows are genotype platform-specific, we considered v5 (GSA-based) and pre-v5 research participants separately. We summed the ancestry calls for each population from the first stage of the algorithm across all the haplotypes inferred to carry the rs200560853 risk variant.

### IBD analysis

For IBD analysis, we computed phasedIBD between the carrier haplotypes against a reference panel composed of 23andMe research participants, genotyped on the v5 (GSA-based) array, who have consented to participate in research and answered survey questions reporting that their four grandparents were all born within one country (excluding the US and Canada). We required exact haplotype matches, setting phasedIBD parameters L_m=100 and L_f=0. Reference panel individuals are counted as an IBD match if they share an IBD segment spanning across the locus with any of the carriers’ haplotypes inferred to carry the allele.

### 4GP allele frequencies estimates

In order to obtain more precise minor allele frequency estimates for rs200560853 in the 4GP cohorts, we imputed the 4GP individuals on a +/- 5.5mb flanking region surrounding the missense variant (GRCh38: chr2:167,000,000 - 178,000,000) using the imputation reference panel described above, following the same imputation procedure as described in the ‘Genotype Imputation’ section. After obtaining their imputation dosages, we estimated the MAF as a sum of imputed dosages divided by twice the number of individuals in each of the 4GP countries/geographic divisions. To obtain the 95% bootstrap confidence interval for the MAFs, we resampled the imputed dosages within each 4GP geographic division 1,000 times with replacement, calculated the resampled MAF for rs200560853, and constructed the 95% CI as the 2.25-th and 97.5-th percentiles of the 1,000 resampled MAF estimates.

### Data Availability

The full GWAS summary statistics for the 23andMe discovery data set will be made available through 23andMe to qualified researchers under an agreement with 23andMe that protects the privacy of the 23andMe participants. Datasets will be made available at no cost for academic use. Please visit https://research.23andme.com/collaborate/#dataset-access/ for more information and to apply to access the data.

## Acknowledgments

The authors would like to thank Patrick Koenig for pymol rendering of protein structures. We thank 23andMe customers who consented to participate in research for enabling this study. We also thank employees of 23andMe who contributed to the development of the infrastructure that made this research possible.

The following members of the 23andMe Research Team contributed to this study: Stella Aslibekyan, Adam Auton, Elizabeth Babalola, Robert K. Bell, Jessica Bielenberg, Jonathan Bowes, Katarzyna Bryc, Ninad S. Chaudhary, Daniella Coker, Sayantan Das, Emily DelloRusso, Sarah L. Elson, Nicholas Eriksson, Teresa Filshtein, Pierre Fontanillas, Will Freyman, Zach Fuller, Chris German, Julie M. Granka, Karl Heilbron, Alejandro Hernandez, Barry Hicks, David A. Hinds, Ethan M. Jewett, Yunxuan Jiang, Katelyn Kukar, Alan Kwong, Yanyu Liang, Keng-Han Lin, Bianca A. Llamas, Matthew H. McIntyre, Steven J. Micheletti, Meghan E. Moreno, Priyanka Nandakumar, Dominique T. Nguyen, Jared O’Connell, Aaron A. Petrakovitz, G. David Poznik, Alexandra Reynoso, Shubham Saini, Morgan Schumacher, Leah Selcer, Anjali J. Shastri, Janie F. Shelton, Jingchunzi Shi, Suyash Shringarpure, Qiaojuan Jane Su, Susana A. Tat, Vinh Tran, Joyce Y. Tung, Xin Wang, Wei Wang, Catherine H. Weldon, Peter Wilton, Corinna D. Wong. J.S., J.O., B.H., W.W., K.B., J.J.B, V.V., W.F., N.S.A., A.A., S.S. are employed by and hold stock or stock options in 23andMe, Inc.

