## Supplementary tables and figures for "GWAS of cataract in Puerto Ricans identifies a novel large-effect variant in ITGA6"

Supplementary Table 1: Age/ Gender characteristics in the cataract GWAS.

Supplementary Table 2: SuSiE 95% credible set of the ITAG6 locus.

Supplementary Table 3: Effect allele frequencies of rs200560853 (23andMe vs gnomAD vs TOPMed)

Supplementary Table 4: The 12 transcript isoforms of ITAG6 (ENSG00000091409) based on the ENSEMBL resource.

Supplementary Figure 1: Effective population size in Puerto Rico

Supplementary Figure 2: Ancestry of inferred carrier individuals' haplotypes on the GSA-based platform (*top*) and the v1-v4 platform (*bottom*) along chromosome 2.

Supplementary Table 1: Age/ Gender characteristics in the cataract GWAS.

| | case | control | Total |
|---|---|---|---|
| Gender | | | |
| Male | 1,155 | 18,449 | 19,604 |
| Female | 1,905 | 23,441 | 24,346 |
| Age | | | |
| (0, 30] | 39 | 8,806 | 8,845 |
| ( 30, 45 ] | 110 | 16,729 | 16,839 |
| ( 45, 60 ] | 398 | 10,906 | 11,304 |
| ( 60, Inf ) | 2,513 | 5,449 | 7,962 |

Supplementary Table 2: SuSiE 95% credible set of the ITAG6 locus. (OR: odds ratio, with respect to the effect allele; CI: confidence interval; EAF: effect allele frequency; PIP: posterior inclusion probability)

| rsID | Cytoband | Position (GRCh38) | Effect Allele | Ref Allele | Gene Context | P-value (LRT) | OR | 95% CI | EAF | PIP |
|---|---|---|---|---|---|---|---|---|---|---|
| rs376241845 | 2q31.1 | 172,406,735 | T | C | DLX2---[]--ITGA6 | $2.14 \times 10^{-11}$ | 11.64 | [5.92, 22.86] | $9.01 \times 10^{-4}$ | 0.083 |
| rs200560853 | 2q31.1 | 172,479,990 | G | C | [ITGA6] | $2.85 \times 10^{-12}$ | 12.67 | [6.49, 24.77] | $8.89 \times 10^{-4}$ | 0.775 |
| rs1046007618 | 2q31.1 | 172,549,415 | A | C | ITGA6--[]-PDK1 | $1.83 \times 10^{-11}$ | 10.52 | [5.53, 20.02] | $9.71 \times 10^{-4}$ | 0.110 |

Supplementary Table 3: Effect allele frequencies of rs200560853 (23andMe vs gnomAD vs TOPMed).
gnomAD: https://gnomad.broadinstitute.org/variant/2-172479990-C-G?dataset=gnomad_r3
TOPMed: https://bravo.sph.umich.edu/freeze8/hg38/variant/snv/2-172479990-C-G

| | 23andMe | gnomAD v3.1.2 | TOPMed Freeze8 |
|---|---|---|---|
| Puerto Rican | $8.89\times10^{-4}$ | NA | NA |
| Latino | $1.88\times10^{-4}$ | $1.97\times10^{-4}$ | $1.4\times10^{-3}$ |
| European | 0 | 0 | 0 |
| African | 0 | 0 | 0 |
| East Asian | 0 | 0 | 0 |
| South Asian | 0 | 0 | 0 |

Supplementary Table 4: The 12 transcript isoforms of ITAG6 (ENSG00000091409) based on the ENSEMBL resource. rs200560853 C>G causes a Cys>Trp amino acid change in all long protein coding transcripts.

| Transcript ID | Name | bp | Protein (aa) | Consequence | Biotype | CCDS | UniProt Match | Flags |
|---|---|---|---|---|---|---|---|---|
| ENST00000684293 | ITGA6-212 | 5816 | 1073 | Cys496Trp | Protein coding | CCDS2249 | P23229-2 | MANE Select; Ensembl Canonical; GENCODE basic; APPRIS ALT1 |
| ENST00000442250 | ITGA6-206 | 5803 | 1130 | Cys535Trp | Protein coding | - | P23229-1 | MANE Plus Clinical; GENCODE basic; TSL:5 |
| ENST00000409080 | ITGA6-202 | 5686 | 1091 | Cys496Trp | Protein coding | CCDS46451 | P23229-3 | GENCODE basic; APPRIS P3; TSL:2 |
| ENST00000264107 | ITGA6-201 | 5488 | 1058 | Cys496Trp | Protein coding | - | A0A8C8KBL6 | GENCODE basic; TSL:1 |
| ENST00000409532 | ITGA6-203 | 3544 | 954 | Cys377Trp | Protein coding | CCDS82534 | P23229-7 | GENCODE basic; TSL:2 |
| ENST00000458358 | ITGA6-207 | 3261 | 1086 | Cys491Trp | Protein coding | - | P23229-5 | GENCODE basic; TSL:5 |
| ENST00000412899 | ITGA6-204 | 970 | 231 | - | Protein coding | - | C9JXX7 | TSL:1; CDS 3' incomplete |
| ENST00000416789 | ITGA6-205 | 853 | 258 | - | Protein coding | - | H7BZ97 | TSL:3; CDS 5' incomplete |
| ENST00000469534 | ITGA6-208 | 581 | No protein | - | Retained intron | - | - | TSL:2 |
| ENST00000475302 | ITGA6-210 | 577 | No protein | - | Retained intron | - | - | TSL:3 |
| ENST00000497107 | ITGA6-211 | 573 | No protein | - | Retained intron | - | - | TSL:2 |
| ENST00000470259 | ITGA6-209 | 365 | No protein | - | Retained intron | - | - | TSL:3 |

Supplementary Figure 1: Effective population size for the Puerto Rican population estimated using IBDNe on 200 individuals. The dark line shows the point estimate for the effective population size, and the gray ribbon shows the 95% confidence interval. The y-axis is on a logarithmic scale.

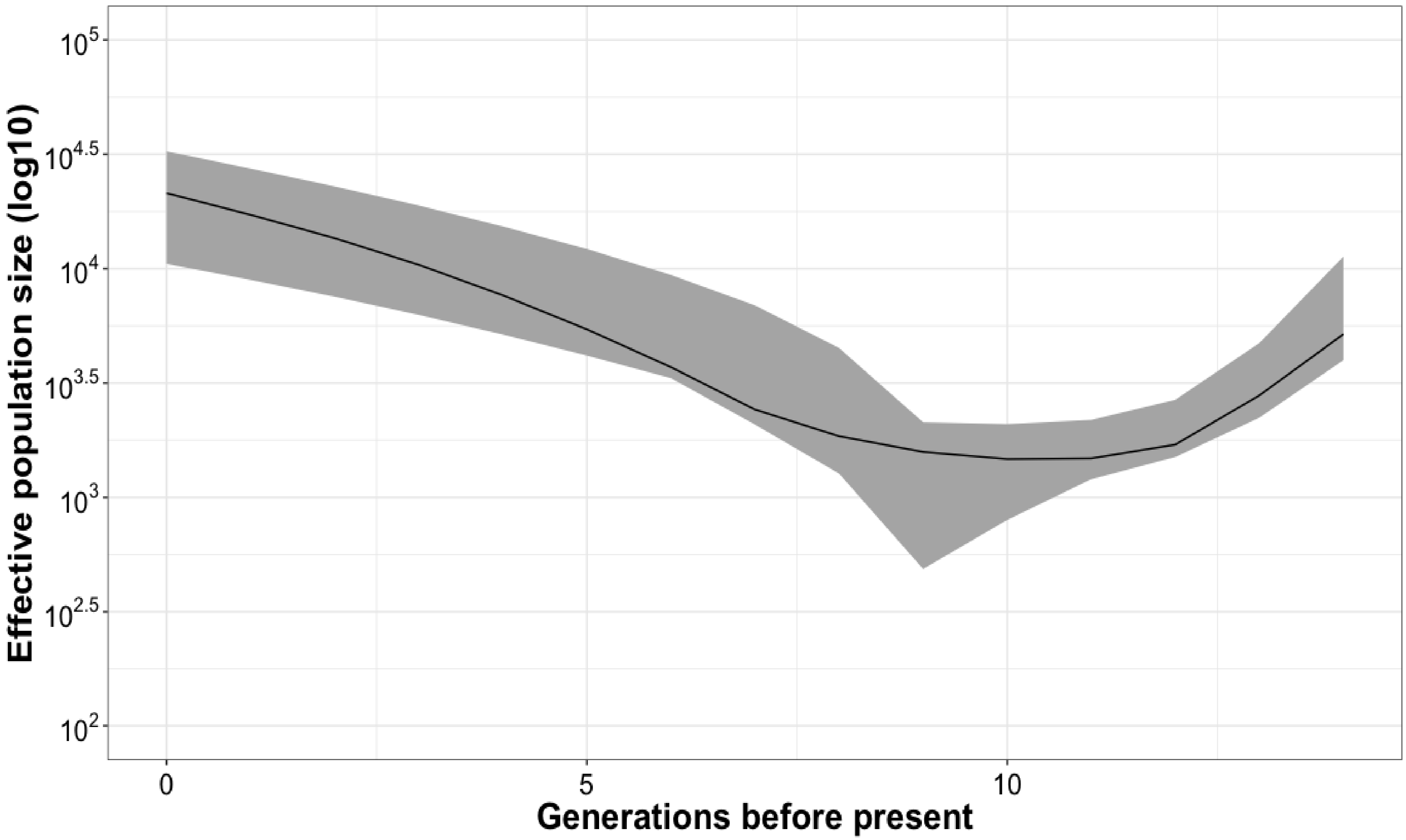

Supplementary Figure 2: Ancestry of inferred carrier individuals' haplotypes on the GSA-based platform (*top*) and the v1-v4 platform (*bottom*) along chromosome 2. Red triangle indicates the location of the rs200560853 missense variant. The x-axis corresponds to the physical position of each 300-SNP Ancestry Composition window, and the y-axis shows the number of windows assigned to be of each ancestry, designated by color. Both platforms show a clear peak of North African ancestry at the window overlapping the locus of interest.

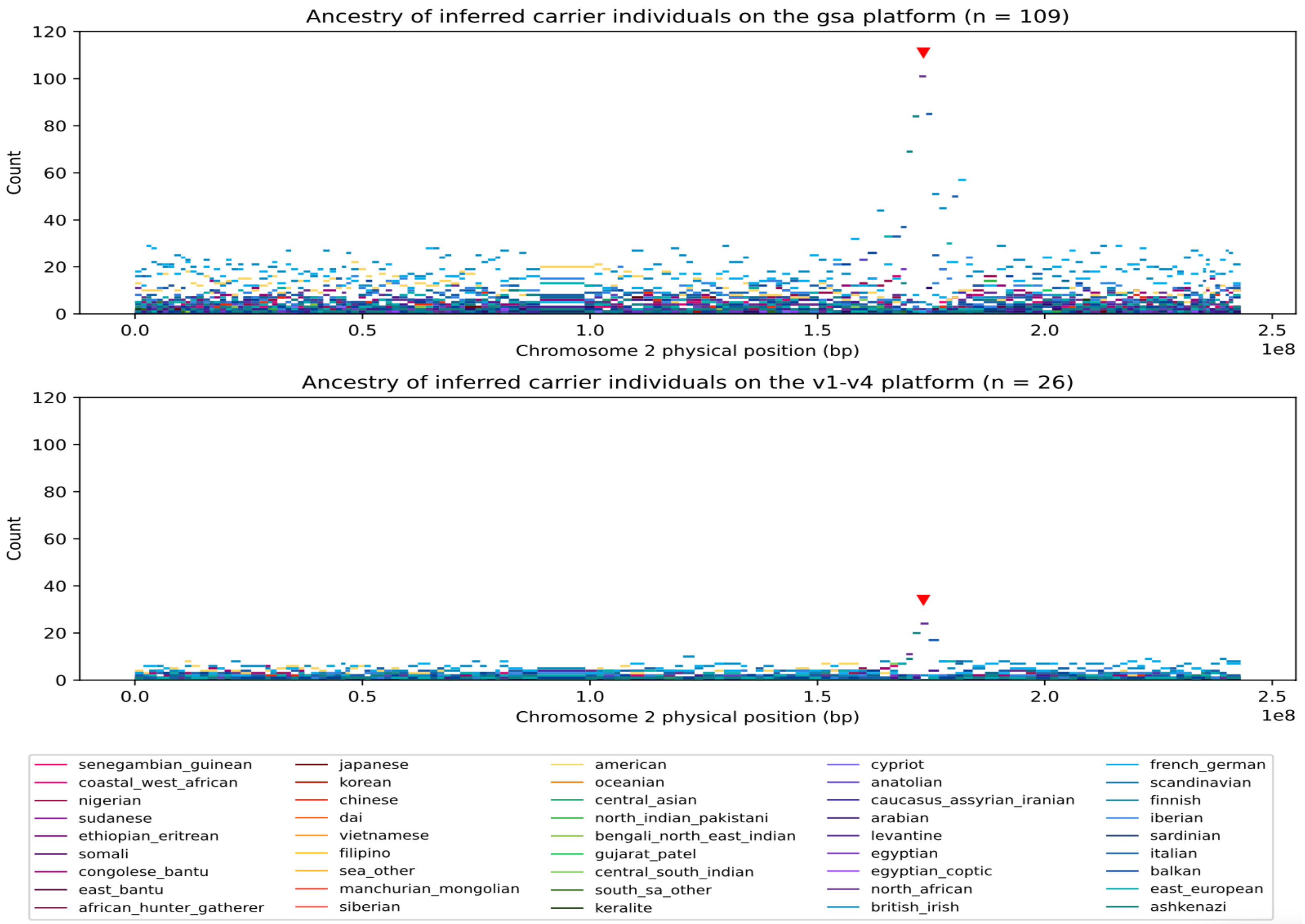
